# Breakthrough SARS-CoV-2 Infection Outcomes in Vaccinated Patients with Chronic Liver Disease and Cirrhosis: A National COVID Cohort Collaborative Study

**DOI:** 10.1101/2022.02.25.22271490

**Authors:** Jin Ge, Jean C. Digitale, Mark J. Pletcher, Jennifer C. Lai, the N3C Consortium

**Author notes:** **<u>Corresponding Author:</u>** Jin Ge, MD, MBA, 513 Parnassus Avenue, S-357, San Francisco, CA 94143.

## Abstract

**Background and Aims:** The incidence and outcomes of breakthrough SARS-CoV-2 infections in vaccinated chronic liver disease (CLD) patients have not been well-characterized in non-veteran populations. We used the National COVID Cohort Collaborative (N3C), a dataset of 10.7 million patients, of whom 0.9 million have vaccination data, to describe outcomes in vaccinated CLD patients.

**Methods:** We identified all CLD patients with or without cirrhosis regardless of vaccination status who had SARS-CoV-2 testing in the N3C Data Enclave as of 1/15/2022. We used Poisson regression to estimate incidence rates of breakthrough infections and Cox survival analyses to associate vaccination status with all-cause mortality at 30 days among infected CLD patients.

**Results:** We isolated 278,457 total CLD patients: 43,079 (15%) vaccinated and 235,378 (85%) unvaccinated. Of the 43,079 vaccinated CLD patients, 32,838 (76%) were without cirrhosis and 10,441 (24%) were with cirrhosis. Estimated incidence rates for breakthrough infections were 5.6 and 5.1 per 1,000 person-months for 27,235 fully vaccinated CLD patients without cirrhosis and for 8,218 fully vaccinated CLD patients with cirrhosis, respectively.

Of the 68,048 unvaccinated and 10,441 vaccinated CLD patients with cirrhosis in our cohort, 15% and 3.7%, respectively, developed SARS-CoV-2 infection. The combined 30-day all-cause rate of mechanical ventilation (without death) or death after SARS-CoV-2 infection for unvaccinated and vaccinated CLD patients with cirrhosis were 15.2% and 7.7%, respectively. Compared to unvaccinated patients with cirrhosis, full vaccination was associated with a 0.34-times adjusted hazard of death at 30 days.

**Conclusions:** In this N3C Data Enclave study, breakthrough infection rates were similar amongst CLD patients with and without cirrhosis. Full vaccination was associated with a 66% reduction in risk of all-cause mortality among CLD patients with cirrhosis after infection. These results provide an additional impetus for increasing vaccination uptake among patients with severe liver disease.

## Introduction

The advent of safe and effective vaccines against Severe Acute Respiratory Syndrome Coronavirus 2 (SARS-CoV-2) has substantially altered the trajectory of the COVID-19 pandemic. These vaccines have been demonstrated to be highly effective in both clinical trials and real-world studies to decrease the risk of severe disease due to SARS-CoV-2 infection. ^2–4^ Nevertheless, despite these encouraging findings, patients with advanced liver disease have well-recognized immune dysfunction with attenuated responses to other vaccines.^5–7^ Recent studies showed that 22.2% of patients with chronic liver diseases (CLD) who received whole virion SARS-Cov-2 vaccines generated no antibody responses and that 24.0% of those who received messenger ribonucleic acid-based (mRNA) SARS-CoV-2 vaccines generated poor [defined as <250U/mL] antibody responses.^8,9^ Moreover, multiple previous studies in various large cohorts have demonstrated that SARS-CoV-2 infection in patients with cirrhosis is associated with higher rates of morbidity and mortality.^10–14^ These data highlight the growing importance of optimizing vaccination strategies for SARS-CoV-2 infection in this vulnerable patient population.^15^

A recent study utilizing the United States Department of Veterans Affairs Clinical Data Warehouse demonstrated that receiving an mRNA based COVID-19 vaccine was associated with a 65% reduction in SARS-CoV-2 infection in veterans with CLD – a substantially lower reduction than that seen in the general population (>95%).^16^ Despite this attenuated clinical response, however, SARS-CoV-2 infection in fully vaccinated veterans with cirrhosis was associated with a 78% reduction in mortality risk compared to unvaccinated veterans with cirrhosis.^17^ While these studies demonstrated the efficacy, albeit attenuated, of vaccination in patients with cirrhosis, the underlying populations were greater than 96% male and greater than 55% non-Hispanic White, consistent with a veteran population.^16,17^

Significant knowledge gaps remain regarding COVID-19 vaccinations in non-veteran populations with cirrhosis. We, therefore, leveraged the National COVID Cohort Collaborative (N3C), which has been previously described in detail,^1,10,18–20^ to address these gaps. As of January 15, 2021, 280 clinical sites had signed data transfer agreements and 69 sites, of which 44 sites had vaccination information, had completed data harmonization into the N3C Data Enclave: a diverse and nationally representative central repository of harmonized electronic health record (EHR) data. We used the N3C Data Enclave to answer two questions relevant to SARS-CoV-2 vaccinations in CLD patients with cirrhosis: Do the rates of breakthrough infection (defined as SARS-CoV-2 infection after vaccination) differ between CLD patients with and without cirrhosis? And, what is the risk reduction in mortality associated with SARS-CoV-2 vaccination among CLD patients with cirrhosis who are infected with SARS-CoV-2?

## Methods

### The National COVID Cohort Collaborative

The N3C is a centralized, curated, harmonized, secure, and nationally representative clinical data resource with embedded analytical capabilities that has been previously described in detail in multiple analyses.^1,10,18–20^ The N3C Data Enclave includes EHR data of patients who were tested for SARS-CoV-2 or had related symptoms after January 1, 2020 with lookback data provided to January 1, 2018. All EHR data in the N3C Data Enclave are harmonized in the Observational Medical Outcomes Partnership (OMOP) common data model, version 5.3.1.^21^ For all analyses, we utilized the de-identified version of the N3C Data Enclave, version 60, dated January 14, 2022 and accessed on January 15, 2022. To protect patient privacy, all dates in the de-identified N3C Data Enclave are uniformly shifted up to ± 180 days within each partner site. Due to the availability of vaccination data in a limited number of sites, we restricted all analyses to only those partner sites with vaccination data.

### Definitions of SARS-CoV-2 Infection Status, Chronic Liver Disease/Cirrhosis, and Laboratory Data

SARS-CoV-2 definitions were based on culture or nucleic acid amplification testing for SARS-CoV-2 as described in previous works.^10^ As the de-identified N3C Data Enclave contains date-shifted data and lacks information on SARS-CoV-2 genomic sequencing, we could not differentiate between variants of SARS-CoV-2. CLD and cirrhosis definitions were based on OMOP identifiers as defined in previous work conducted with the N3C Data Enclave.^10^ All patients who had undergone orthotopic liver transplantation were excluded from all analyses.^10^ As dates are uniformly shifted within each partner site in the de-identified N3C Data Enclave, we calculated a “maximum data date” to reflect the last known date of records for each site. We used this “maximum data date” to exclude patients who were vaccinated or tested for SARS-Cov-2 within 90 days of this date to allow for adequate follow-up time and account for potential delays in data harmonization.

### Definition of Vaccination Status

Vaccinations were defined based on OMOP drug exposure concepts delineated in vaccination studies on immunocompromised patients utilizing the N3C Data Enclave and are provided in Supplemental Table 1.^19^ Of note, we only included OMOP concept identifiers for the three SARS-CoV-2 vaccination regimens with Food and Drug Administration emergency use authorizations or approvals in the United States: two mRNA vaccines from Pfizer-BioNTech (BNT162b2) and Moderna (mRNA-1273), and one viral vector vaccine from Johnson & Johnson/Janssen (JNJ-784336725).^2–4^ All vaccines not authorized for use in the United States were excluded.

We defined “partial vaccination” as starting on day 7 after one dose of any mRNA vaccine, and we defined “full vaccination” as 14 days after the completion of the initial recommended dosing regimen for any vaccine (two doses for mRNA vaccines or one dose for viral vector vaccine). Under this commonly used definition for “fully vaccinated,” one dose of viral vector vaccine is equivalent to two doses of mRNA vaccines and regarded as two doses in all numeric counts of vaccination doses in summary statistics.^22^ Due to data limitations, we treated patients who received additional (“booster”) doses administered over and above full vaccination regimens as only fully vaccinated, and where possible, separately examined the effect of number of doses received. “Breakthrough infections” were defined as SARS-CoV-2 infections in partially vaccinated or fully vaccinated patients with no history of SARS-CoV-2 infection prior to vaccination.

We defined “unvaccinated” patients as those who did not have an associated OMOP drug exposure concept identifier for vaccination defined above (Supplemental Table 1) – as such this definition also includes patients whose vaccination status is not known within the N3C Data Enclave. In addition, patients who had acquired SARS-CoV-2 infection prior to seven days after one dose of any mRNA vaccine or 14 days after one dose of viral vector vaccine were also considered to be unvaccinated in our analyses. Patients who develop an infection prior to day 7 after one dose of any mRNA vaccine or day 14 after one dose of viral vector vaccine, therefore, do not contribute any follow-up time to partial and full vaccination statuses.

### Study Design and Questions of Interest

Using definitions for SARS-CoV-2 positivity, CLD and cirrhosis, and vaccination status, we isolated our adult patient (documented age ≥ 18 years) study populations for two study questions: 1. What is the relative rate of breakthrough SARS-CoV-2 infection comparing patients with and without cirrhosis who are at least partially vaccinated with no history of SARS-CoV-2 infection prior to vaccination? In this question, we sought to compare the rates of breakthrough SARS-CoV-2 infection in vaccinated CLD patients with cirrhosis versus vaccinated CLD patients without cirrhosis. 2. What is the reduction in mortality associated with vaccination among CLD patients with cirrhosis who are infected with SARS-CoV-2? In this question, we compared all-cause mortality at 30 days in (partially and fully) vaccinated CLD patients with cirrhosis who acquire breakthrough SARS-CoV-2 infection versus unvaccinated CLD patients with cirrhosis who acquire SARS-CoV-2 infection.

### Outcomes

The primary outcomes in our study were breakthrough SARS-CoV-2 infection and all-cause mortality at 30 days after SARS-CoV-2 infection. Secondary outcomes included hospitalization and mechanical ventilation within 30 days. As in previous works, the outcomes of death, mechanical ventilation, and hospitalization are centrally defined based on N3C shared logic.^1,10,18^

### Baseline Characteristics

Baseline demographic characteristics extracted from N3C Data Enclave included age, sex, race/ethnicity, height, weight, body mass index, and state of origin. States were classified into four geographic regions (Northeast, Midwest, South, and West) defined by the Center for Disease Control and Prevention’s (CDC) National Respiratory and Enteric Virus Surveillance System (NREVSS).^23^ Patients were categorized as living in “Other/Unknown” region if they originated from territories not otherwise classified or if state of origin was not available. We utilized a “Modified Charlson Index” based on the Charlson Comorbidity Index excluding “mild liver disease” and “severe liver disease” as defined in previous analyses utilizing N3C.^10^

Components of common laboratory tests (basic metabolic panel, complete blood count, liver function tests, and serum albumin) were extracted based on N3C shared logic sets except for international normalized ratio (INR), which was based on previously defined concept sets.^10^ We extracted the most number of distinct laboratory values to calculate the model for end-stage liver disease-sodium (MELD-Na) score closest to or on the index date from within 30 days before to 7 days after the date of the earliest positive test (for SARS-CoV-2 infected patients) or negative test (for non-infected patients). Over half (56%) of patients had laboratory tests within 2 days of the date of the earliest positive or negative test. Of note, approximately 16% of CLD patients with cirrhosis in the eligible population had full laboratory data to calculate MELD-Na scores.

### Statistical Analyses

Clinical characteristics and laboratory data were summarized by medians and interquartile ranges (IQR) for continuous variables or numbers and percentages (%) for categorical variables. Comparisons between groups were performed using Kruskal-Wallis and chi-square tests where appropriate. All patients were followed until their last recorded visit occurrence, procedure, measurement, observation, or condition occurrence in the N3C Data Enclave.

For our first analysis of breakthrough infections between vaccinated CLD patients with cirrhosis versus those without cirrhosis, we estimated unadjusted incidence rates and adjusted incidence rate ratios (IRRs) using Poisson regression models implemented as generalized estimating equations with robust standard errors (SEs).^24^ We created three distinct adjusted Poisson regression models to evaluate controlling for: 1) partial versus full vaccination status, 2) initial vaccination type (BNT162b2, mRNA-1273, or JNJ-784336725) in addition to partial versus full status, and 3) the number of vaccine doses (0-4). All multivariable Poisson regression models accounted for age, sex, race/ethnicity, CLD etiology, modified Charlson scores, and region of origin.

At-risk person-time for incidence rate estimation accrued from 7 days after the first dose of mRNA vaccines and from 14 days after the first dose of viral vector vaccines to the date of breakthrough infection, death, or last available visit occurrence date in the N3C Data Enclave (censoring). Participants contributed person-time to partial vaccination status from seven days after the first dose of mRNA vaccine to the date of the second dose, breakthrough infection, death, or censoring in the unadjusted Poisson regression model for breakthrough infections in partially vaccinated patients. Participants contributed person-time to full vaccination status from 14 days after the second dose of mRNA vaccine or 14 days after the first dose of viral vector vaccine to breakthrough infection, death, or censoring in the unadjusted Poisson regression for breakthrough infections in fully vaccinated patients. Of note, under these definitions, participants who received viral vector vaccines do not accumulate person-time for partial vaccination status. For adjusted Poisson regression models evaluating all (partially and fully) vaccinated patients, participants’ person-times are sums of contributions to both partial vaccination and full vaccination statuses.

For the second analysis of all-cause mortality in vaccinated CLD patients with cirrhosis with breakthrough SARS-CoV-2 infections compared to unvaccinated CLD patients with cirrhosis and SARS-CoV-2 infections, we used Cox proportional hazard models to evaluate the associations between vaccination status (partial or full vs. unvaccinated) and mortality among patients with cirrhosis who tested positive for SARS-CoV-2.^25^ At risk person-time for this analysis accrued from the index date of positive SARS-CoV-2 test to death or last available visit occurrence date in the N3C Data Enclave (censoring). Cox regression models were adjusted for age, sex, race/ethnicity, CLD etiology, modified Charlson score, and region of origin.

Two-sided p-values < 0.05 were considered statistically significant in all analyses. Data queries, extractions, and transformations of OMOP data elements and concepts in the N3C Data Enclave were conducted using the Palantir Foundry implementations of Spark-Python, version 3.6, and Spark-SQL, version 3.0. Statistical analyses were performed using the Palantir Foundry implementation of Spark-R, version 3.5.1 “Feather Spray” (R Core Team, Vienna, Austria).

### Institutional Review Board Oversight

Submission of data from individual centers to N3C are governed by a central institutional review board (IRB) protocol #IRB00249128 hosted at Johns Hopkins University School of Medicine via the SMART IRB40 Master Common Reciprocal reliance agreement. This central IRB covers data contributions and transfer to N3C but does not cover research using N3C data. If elected, individual sites may choose to exercise their own local IRB agreements instead of utilizing the central IRB. As the National Institutes of Health (NIH) National Center for Advancing Translational Sciences (NCATS) is the steward of the repository, data received and hosted by NCATS on the N3C Data Enclave, its maintenance, and its storage are covered under a central NIH IRB protocol to make EHR-derived data available for the clinical and research community to use for studying COVID-19. Our institution has an active data transfer agreement with N3C. This specific analysis of the N3C Data Enclave was approved by N3C under Data Use Agreements titled “[RP-7C5E62] COVID-19 Outcomes in Patients with Cirrhosis” and “[RP-E77B79] COVID-19 Outcomes in Vaccinated Patients with Liver Diseases.” The use of N3C data for this study was authorized by the IRB at the University of California, San Francisco under #21-35861.

## Results

As of January 15, 2022, 69 sites that had completed data transfer were harmonized and integrated into the N3C Enclave, of which 44 sites provided vaccination information. Of the unique patients who had undergone at least one SARS-CoV-2 test, an eligible population of 278,457 CLD patients with or without cirrhosis regardless of vaccination status was assembled, after applying exclusion criteria for transplant status, age, and date shifting in the N3C Enclave. Patient flow and analytic samples for each of the study questions are highlighted in Figure 1.

**Figure 1.**
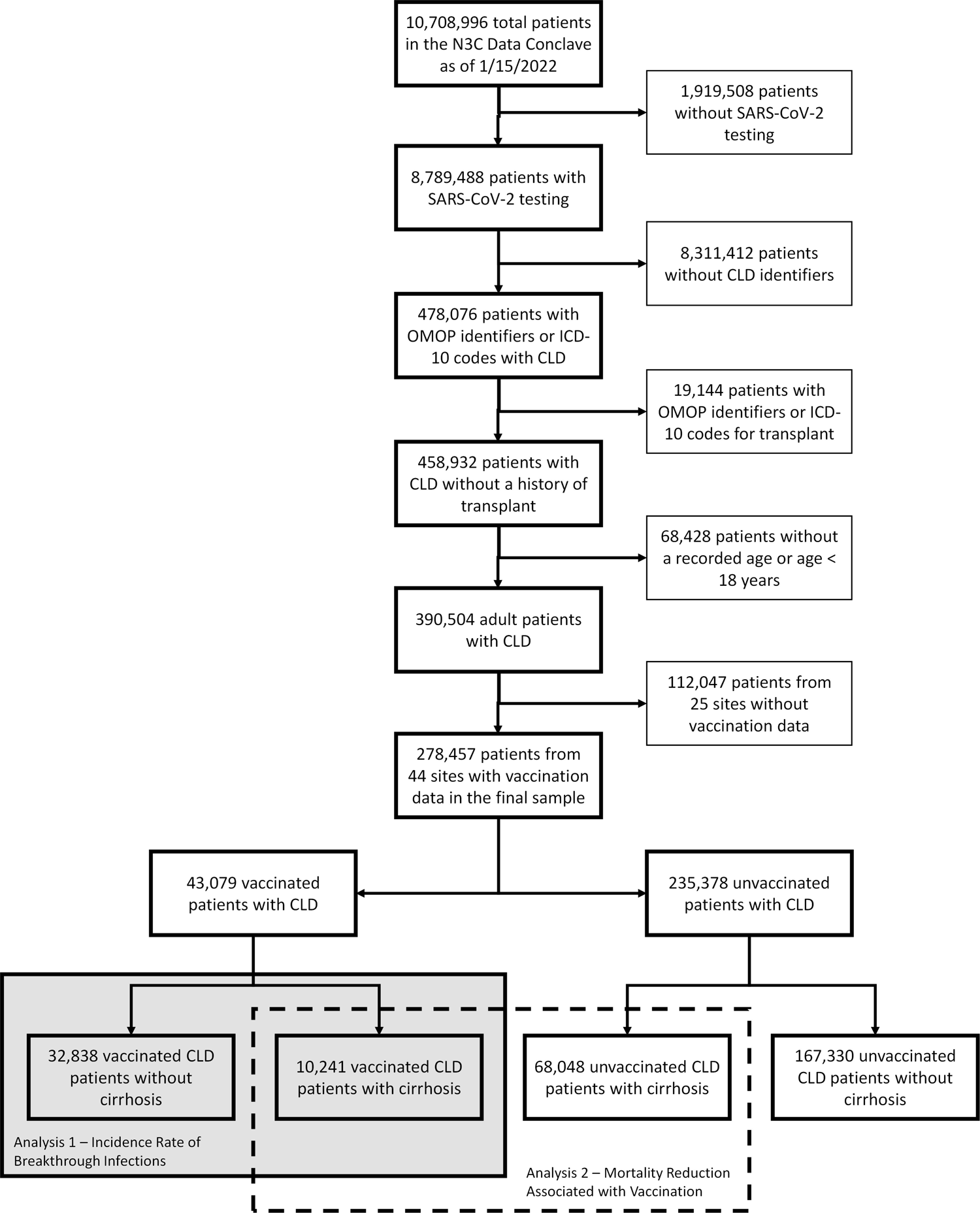
Flowchart of isolation of CLD patients with and without cirrhosis from the main N3C cohort.

### Characteristics of Vaccinated CLD Patients

Based on our vaccination definitions, we isolated 43,079 CLD patients who had at least one dose of SARS-CoV-2 vaccine: this represented 15.8% of the eligible population. Of these 43,079 vaccinated CLD patients, 32,838 (76.2%) did not have cirrhosis and 10,241 (23.8%) had cirrhosis. 95.5% (31,344) of vaccinated CLD patients without cirrhosis and 94.8% (9,711) of vaccinated CLD patients with cirrhosis received a mRNA vaccine as their first dose. 82.9% (27,235) of vaccinated CLD patients without cirrhosis and 80.3% (8,218) of vaccinated CLD patients with cirrhosis completed their initial vaccination series and were fully vaccinated. The baseline characteristics of these patients are presented in Table 1.

**Table 1.** Baseline demographic, clinical, and laboratory characteristics of the 43,079 vaccinated patients with chronic liver diseases with and without cirrhosis.

|  | **Vaccinated CLD without**<br>**Cirrhosis**<br>**(n = 32,838)** | **Vaccinated CLD with**<br>**Cirrhosis**<br>**(n = 10,241)** | **P-Value** |
| --- | --- | --- | --- |
| **Female** | 17,909 (55) | 4,743 (46) | <0.01 |
| **Age (years)** | 57 (45-66) | 61 (53-69) | <0.01 |
| 18-29 | 1,471 (4) | 213 (2) |  |
| 30-49 | 9,135 (28) | 1,765 (17) |  |
| 50-64 | 12,826 (39) | 4,256 (42) |  |
| 65+ | 9,406 (29) | 4,007 (39) |  |
| **Race/Ethnicity** |  |  | <0.01 |
| White | 16,528 (50) | 5,690 (56) |  |
| Black/African-American | 5,073 (15) | 1,725 (17) |  |
| Hispanic | 6,938 (21) | 1,851 (18) |  |
| Asian | 1,931 (6) | 334 (3) |  |
| Unknown/Other | 2,368 (7) | 641 (6) |  |
| **Height (cm)** | 168 (160-175) | 170 (163-178) | <0.01 |
| **Weight (kg)** | 88 (73-104) | 83 (69-100) | <0.01 |
| **BMI (kg/m2)** | 31 (27-36) | 29 (25-34) | <0.01 |
| **Liver Disease Etiology** |  |  | <0.01 |
| NAFLD | 22,817 (69) | 3,549 (35) |  |
| Hepatitis C | 6,230 (19) | 2,036 (20) |  |
| AALD | 1,205 (4) | 2,995 (29) |  |
| Hepatitis B | 1,998 (6) | 514 (5) |  |
| Cholestatic | 159 (0) | 697 (7) |  |
| Autoimmune | 429 (1) | 450 (4) |  |
| **Decompensated Cirrhosis** | 0 (0) | 6,287 (61) |  |
| **Modified Charlson Index†** | 1 (0-3) | 2 (1-5) | <0.01 |
| **Region** |  |  | <0.01 |
| Northeast | 3,981 (12) | 910 (9) |  |
| Midwest | 5,254 (16) | 2,155 (21) |  |
| South | 5,071 (15) | 1,915 (19) |  |
| West | 7,261 (22) | 2,090 (20) |  |
| Other | 11,271 (34) | 3,171 (31) |  |
| **MELD-Na‡** | 9 (7-12) | 14 (10-20) | <0.01 |
| **Initial Vaccine Type** |  |  | <0.01 |
| BNT162b2 | 21,040 (64) | 6,909 (67) |  |
| mRNA-1273 | 10,304 (31) | 2,802 (27) |  |
| JNJ-784336725 | 1,494 (5) | 530 (5) |  |
| **Total Number of Doses** |  |  | <0.01 |
| 1 | 5,603 (17) | 2,023 (20) |  |
| 2 | 20,923 (64) | 6,418 (63) |  |
| 3+ | 6,312 (19) | 1,800 (18) |  |
| **Breakthrough COVID Cases** | 1,290 (4) | 379 (4) | 0.31 |

**Note:** Continuous variables are described as medians with interquartile ranges in parentheses, ordinal and categorical variables are described as counts with percentages in parentheses.
†Modified Charlson Index was calculated based on the original Charlson Comorbidity Score excluding weights for “mild liver disease” and “severe liver disease.”
‡MELD-Na scores were calculated for approximately 16% of the total sample.
Abbreviations: AALD, alcohol-associated liver disease; MELD-Na, Model for End-Stage Liver Disease-Sodium; NAFLD, non-alcoholic fatty liver disease.

In general, compared with vaccinated CLD patients without cirrhosis, those vaccinated with cirrhosis were more likely to be older (median 61 versus 57 years), non-Hispanic White (56% versus 50%), have alcohol related liver disease as their etiology (29% versus 4%), and have higher modified Charlson scores (median 2 versus 1). Vaccinated patients with cirrhosis were also less likely to be female (46% versus 55%) compared to vaccinated patients without cirrhosis. Geographic distributions for the two populations also varied with greater proportions of vaccinated patients with cirrhosis in the Midwest and South.

### Breakthrough Infection in Vaccinated CLD Patients

Unadjusted incidence rates of breakthrough infections in vaccinated patients are presented in Table 2. Among 27,235 fully vaccinated patients without cirrhosis, 979 breakthrough infections occurred for an estimated incidence rate of 5.6 (95% confidence interval [CI] 5.2-5.9) per 1,000 person-months. Among 8,218 fully vaccinated patients with cirrhosis, 269 breakthrough infections occurred for an estimated incidence rate of 5.1 (95% CI 4.5-5.7) per 1,000 person-months.

**Table 2.** Unadjusted incidence rates of breakthrough SARS-CoV-2 infections amongst vaccinated CLD patients.

| Patient Group | Total Person-Months | # Breakthrough Cases | Incidence Rate per 1000 Person-Months (95% CI) |
| --- | --- | --- | --- |
| **Partial Vaccination – Overall** | 61,344 | 421 | 6.5 (5.9-7.2) |
| CLD without Cirrhosis | 46,353 | 311 | 6.4 (5.7-7.1) |
| CLD with Cirrhosis | 149,91 | 110 | 7.1 (5.9-8.6) |
| **Full Vaccination – Overall** | 228,291 | 1248 | 5.5 (5.2-5.8) |
| CLD without Cirrhosis | 175,360 | 979 | 5.6 (5.2-5.9) |
| CLD with Cirrhosis | 52,931 | 269 | 5.1 (4.5-5.7) |
| **Note: Estimated incidence rates were based on unadjusted Poisson regression models with robust standard errors.** |  |  |  |

Adjusted incidence rate ratios estimated from multivariable models including 1) partial versus full vaccination status, 2) initial vaccination type, and 3) number of vaccine doses for all vaccinated patients with and without cirrhosis are presented in Table 3. Of note, the presence of cirrhosis was not associated with a higher risk of breakthrough infection when compared to those without cirrhosis in any model. Full vaccination status, defined as two or more vaccine doses, was associated with a 73% reduced risk for breakthrough infections compared with partial vaccination status. There was a dose-dependent relationship between the number of vaccine doses and reduced risk of breakthrough infections. Moreover, compared with CLD patients who initially received BNT162b2, those who received mRNA-1273 had a 18% reduced risk for breakthrough infections, while those who received JNJ-784336725 had a 54% increased risk. Sex, age, and race/ethnicity were not significantly associated with breakthrough infections.

**Table 3.**
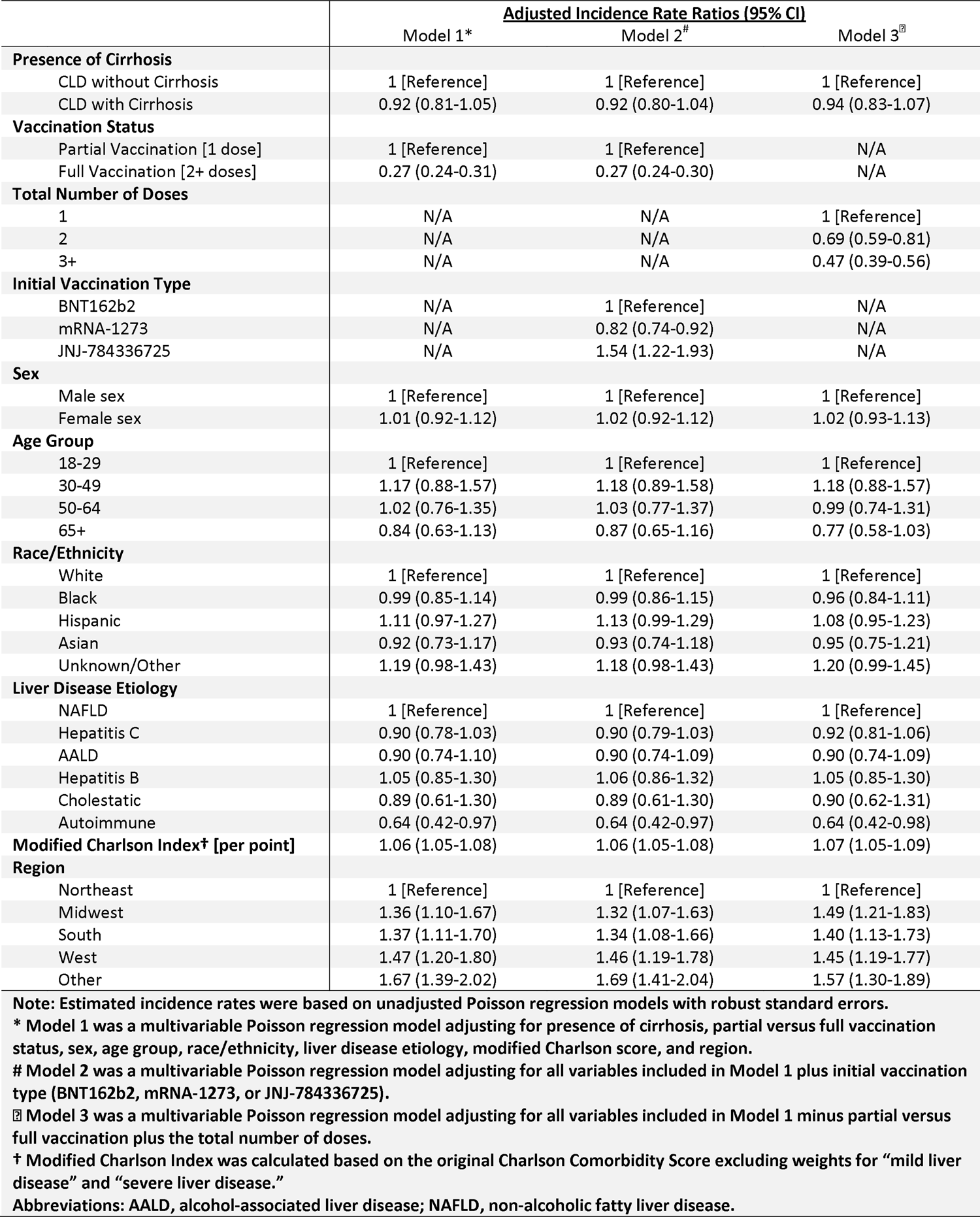
Adjusted incidence rate ratios of clinical and demographic factors associated with breakthrough SARS-CoV-2 infections.

### Characteristics of Unvaccinated versus Vaccinated Patients with Cirrhosis and Infected with SARS-CoV-2

Out of the 68,048 unvaccinated CLD patients with cirrhosis, 10,441 (15.3%) tested positive for SARS-CoV-2. Out of the 10,241 vaccinated CLD patients with cirrhosis (without previous SARS-CoV-2 infections prior to vaccination), 379 (3.7%) had a breakthrough SARS-CoV-2 infection. The baseline demographic and clinical characteristics of the 10,441 unvaccinated CLD patients with cirrhosis infected with SARS-CoV-2 and 379 vaccinated CLD patients with cirrhosis and breakthrough SARS-CoV-2 infections are presented in Table 4. Compared with unvaccinated patients with cirrhosis and SARS-CoV-2 infection, those vaccinated were older (median age 62 years versus 59 years), taller (median height 170cm versus 168cm), and more likely to have other comorbid conditions (median modified Charlson score of 4 versus 3). Vaccinated patients with cirrhosis and breakthrough SARS-CoV-2 infections were less likely to have decompensated cirrhosis (64% versus 70%) compared with unvaccinated patients with cirrhosis and SARS-CoV-2 infections. Of note, there were no significant differences between vaccinated and unvaccinated patients with SARS-CoV-2 infections with regard to race/ethnicity, weight, body mass index, and etiology of liver disease.

**Table 4.** Baseline demographic, clinical, and laboratory characteristics of patients with cirrhosis and SARS-CoV-2 infection.

|  | <u>Unvaccinated Cirrhosis</u><br><u>with SARS-CoV-2</u><br>(n = 10,441) | <u>Vaccinated Cirrhosis with</u><br><u>Breakthrough SARS-CoV-2</u><br>(n = 379) | <u>P-Value</u> |
| --- | --- | --- | --- |
| **Female** | 4,671 (45) | 183 (48) | 0.19 |
| **Age (years)** | 59 (49-67) | 62 (53-70) | <0.01 |
| 18-49* | 2,623 (25) | 70 (18) |  |
| 50-64 | 4,359 (42) | 150 (40) |  |
| 65+ | 3,459 (33) | 159 (42) |  |
| **Race/Ethnicity** |  |  | 0.22 |
| White | 5,569 (53) | 198 (52) |  |
| Black/African-American | 1,742 (17) | 65 (17) |  |
| Hispanic | 2,130 (20) | 67 (18) |  |
| Asian and Unknown/Other* | 1,000 (10) | 49 (13) |  |
| **Height (cm)** | 168 (160-178) | 170 (163-178) | 0.03 |
| **Weight (kg)** | 84 (70-103) | 87 (69-106) | 0.48 |
| **BMI (kg/m2)** | 30 (25-35) | 30 (25-36) | 0.80 |
| **Liver Disease Etiology** |  |  | 0.29 |
| NAFLD | 4,324 (41) | 144 (38) |  |
| Hepatitis C | 2,041 (20) | 78 (21) |  |
| AALD | 2,711 (26) | 95 (25) |  |
| Hepatitis B | 475 (5) | 25 (7) |  |
| Cholestatic/Autoimmune* | 890 (9) | 37 (10) |  |
| **Decompensated Cirrhosis** | 7,283 (70) | 240 (63) | <0.01 |
| **Modified Charlson Index†** | 3 (1-6) | 4 (1-7) | <0.01 |
| **Region** |  |  | <0.01 |
| Northeast | 612 (6) | 21 (6) |  |
| Midwest | 2,317 (22) | 77 (20) |  |
| South | 1,705 (16) | 76 (20) |  |
| West | 863 (8) | 66 (17) |  |
| Other | 4,944 (47) | 139 (37) |  |
| **MELD-Na☒** | 17 (11-24) | 14 (10-18) | 0.01 |
| **Initial Vaccine Type** |  |  |  |
| BNT162b2 |  | 254 (67) |  |
| mRNA-1273 |  | 108 (28) |  |
| **Total Number of Doses** |  |  |  |
| 1 |  | 110 (29) |  |
| 2+* |  | 269 (71) |  |

**Note:** Continuous variables are described as medians with interquartile ranges in parathesis, ordinal and categorical variables are described as values with percentages in parathesis.
\* N3C policy requires all cells that contain fewer than 20 persons to be reported as <20 or collapsed with other categories
† Modified Charlson Index was calculated based on the original Charlson Comorbidity Score excluding weights for “mild liver disease” and “severe liver disease.”
☒ MELD-Na scores were calculated for approximately 16% of the total sample.
Abbreviations: AALD, alcohol-associated liver disease; MELD-Na, Model for End-Stage Liver Disease-Sodium; NAFLD, non-alcoholic fatty liver disease.

Full MELD-Na components were available in 1,689 patients and serum albumin was available in 4,996 patients, these figures respectively represented 15.6% and 46.2% of patients with both cirrhosis and SARS-CoV-2 infection. Among unvaccinated cirrhosis patients with SARS-CoV-2 infections, the median (IQR) MELD-Na was 17 (11-24) and median (IQR) serum albumin was 3.2 g/dL (2.6-3.8 g/dL). Among vaccinated cirrhosis patients with breakthrough SARS-CoV-2 infections, the median (IQR) MELD-Na was 14 (10-18) and median (IQR) serum albumin was 3.3 g/dL (2.8-3.9 g/dL). Patient outcomes among unvaccinated and vaccinated patients with cirrhosis and SARS-CoV-2 infection are shown in Figure 2. The overall all-cause 30-day mechanical ventilation (without death) and death rates after SARS-CoV-2 infection were 6.2% and 9.0%, respectively, among unvaccinated patients. The combined 30-day ventilation or death rate for vaccinated CLD patients with cirrhosis was 7.7% (N3C policy limits reporting of cells and figures with fewer than 20 persons, therefore we combined persons who experienced mechanical ventilation (without death) or death among vaccinated CLD patients with cirrhosis who had breakthrough SARS-CoV-2).

**Figure 2.**
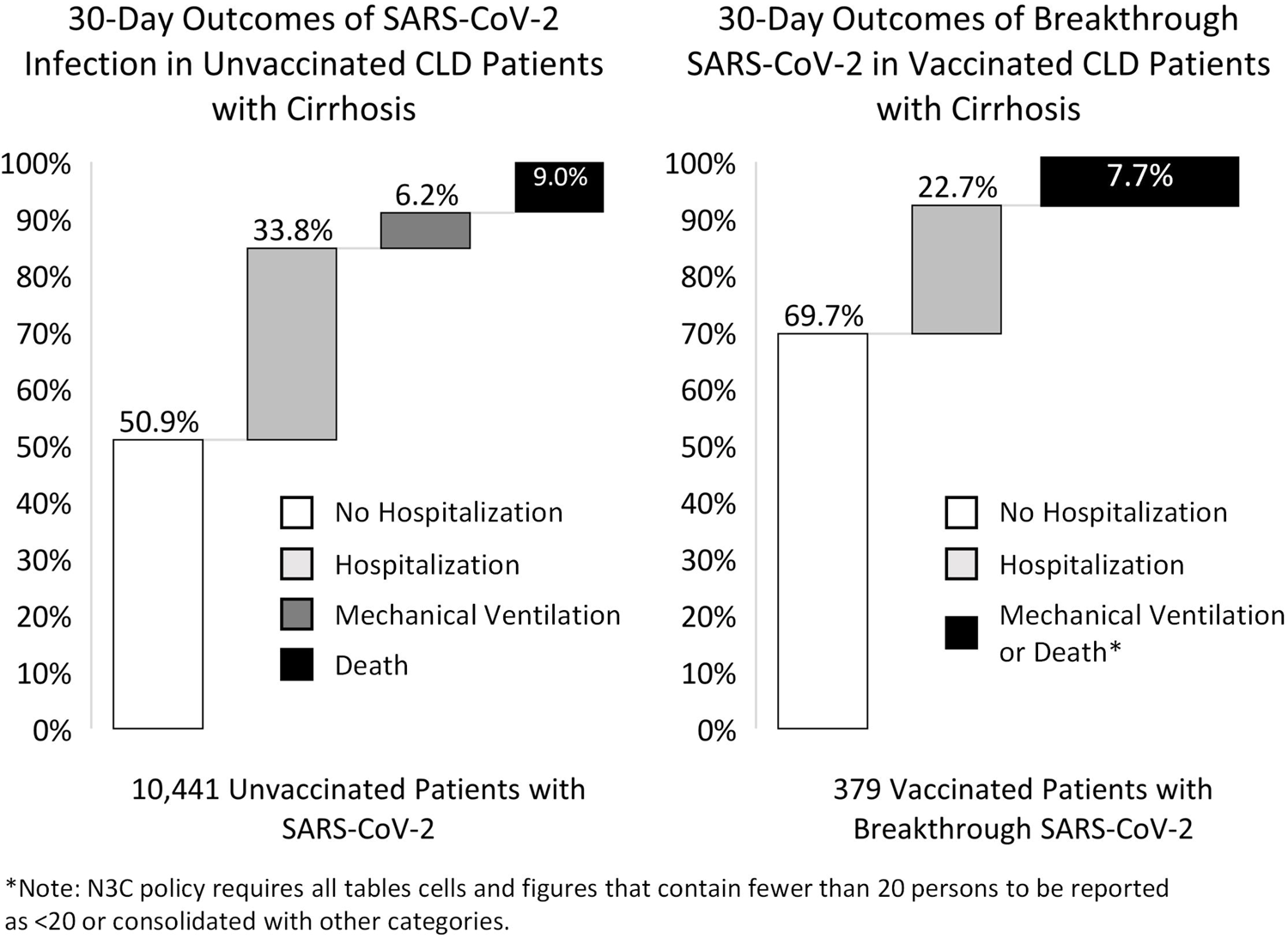
30-day outcomes of SARS-CoV-2 infection in unvaccinated and vaccinated CLD patients with cirrhosis.

### Associations between Vaccination and Death in Patients with Cirrhosis Infected with SARS-CoV-2

Demographic and clinical factors associated with all-cause 30-day mortality among CLD patients with cirrhosis who were infected with SARS-CoV-2 are presented in Table 5. In univariable Cox regression analyses, partial vaccination was not significantly associated with a reduction in all-cause mortality at 30 days (Hazard Ratio [HR] 0.85, 95% CI 0.43-1.72, p = 0.67) while full vaccination was significantly associated with a 0.39-times hazard of death within 30 days (HR 0.39, 95% CI 0.19-0.78, p < 0.01). In adjusted Cox regression analyses, partial vaccination was again not significantly associated with a reduction in all-cause mortality at 30 days (adjusted Hazard Ratio [aHR] 0.78, 95% CI 0.39-1.57, p = 0.49). Full vaccination, however, was associated with lower hazard of death within 30 days (aHR 0.34, 95% CI 0.17-0.68, p < 0.01) in multivariable analyses. Of note, chronic hepatitis B as etiology (aHR 0.67, 95% CI 0.47-0.95, p = 0.03), cholestatic liver diseases as etiology (aHR 0.37, 95% CI 0.24-0.57, p < 0.01), and location in the Midwest (aHR 0.72, 95 CI 0.55-0.96, p=0.02) were associated with lower 30-day mortality hazards in multivariable analyses. Every point increase in modified Charlson score (aHR 1.03, 95% CI 1.01-1.05, p < 0.01) was associated with higher 30-day mortality hazards in multivariable analyses.

**Table 5.** Associations of vaccination status with all-cause 30-day mortality in patients with cirrhosis and infected with SARS-CoV-2.

|  | Univariable Cox Regression |  |  | Multivariable Cox Regression |  |  |
| --- | --- | --- | --- | --- | --- | --- |
|  | HR | 95% CI | P-Value | aHR | 95% CI | P-Value |
| **Vaccination Status** |  |  |  |  |  |  |
| Partial Vaccination | 0.86 | 0.43-1.72 | 0.67 | 0.78 | 0.39-1.57 | 0.49 |
| Full Vaccination | 0.39 | 0.19-0.78 | <0.01 | 0.34 | 0.17-0.68 | <0.01 |
| **Age (years)** | 1.03 | 1.03-1.04 | <0.01 | 1.03 | 1.03-1.04 | <0.01 |
| **Female** | 0.85 | 0.75-0.97 | 0.02 | 0.87 | 0.76-0.99 | 0.04 |
| **Race/Ethnicity, no. (%)** |  |  |  |  |  |  |
| White | 1 [Reference] |  |  | 1 [Reference] |  |  |
| Black/African-American | 0.88 | 0.73-1.06 | 0.17 | 0.85 | 0.71-1.03 | 0.11 |
| Hispanic | 0.93 | 0.79-1.10 | 0.38 | 1.02 | 0.86-1.21 | 0.83 |
| Asian | 1.07 | 0.73-1.56 | 0.74 | 1.17 | 0.79-1.73 | 0.42 |
| Unknown/Other | 1.08 | 0.85-1.38 | 0.53 | 1.14 | 0.89-1.46 | 0.30 |
| **Etiology of Liver Disease, no. (%)** |  |  |  |  |  |  |
| NAFLD | 1 [Reference] |  |  | 1 [Reference] |  |  |
| Hepatitis C | 0.90 | 0.76-1.07 | 0.23 | 0.87 | 0.72-1.04 | 0.11 |
| AALD | 0.88 | 0.75-1.03 | 0.11 | 0.95 | 0.80-1.12 | 0.51 |
| Hepatitis B | 0.72 | 0.51-1.01 | 0.06 | 0.67 | 0.47-0.95 | 0.03 |
| Cholestatic | 0.37 | 0.24-0.57 | <0.01 | 0.37 | 0.24-0.57 | <0.01 |
| Autoimmune | 0.88 | 0.60-1.28 | 0.50 | 0.98 | 0.67-1.43 | 0.93 |
| **Modified Charlson Index† [per point]** | 1.05 | 1.04-1.07 | <0.01 | 1.03 | 1.01-1.05 | <0.01 |
| **Region** |  |  |  |  |  |  |
| Northeast | 1 [Reference] |  |  | 1 [Reference] |  |  |
| Midwest | 0.66 | 0.50-0.86 | <0.01 | 0.72 | 0.55-0.96 | 0.02 |
| South | 0.85 | 0.65-1.11 | 0.24 | 0.94 | 0.70-1.24 | 0.65 |
| West | 0.61 | 0.44-0.85 | <0.01 | 0.75 | 0.54-1.05 | 0.10 |
| Other | 0.76 | 0.59-0.97 | 0.03 | 0.81 | 0.63-1.04 | 0.09 |

**Note:** Multivariable Cox regression model was adjusted for sex, age, race/ethnicity, liver disease etiology, modified Charlson score, and region.
† Modified Charlson Index was calculated based on the original Charlson Comorbidity Score excluding weights for “mild liver disease” and “severe liver disease.”
Abbreviations: AALD, alcohol-associated liver disease; aHR, adjusted hazard ratio; CI, confidence interval; HR, hazard ratio; NAFLD, non-alcoholic fatty liver disease.

## Discussion

In this retrospective study of CLD patients in the National COVID Cohort Collaborative, we found that only 15.8% of CLD patients were vaccinated. Nearly all (95%) of vaccinated patients received an mRNA vaccine as the initial dose and 82% were fully vaccinated. Given that CLD patients have been previously demonstrated to have lower immunologic responses to various vaccinations, including to SARS-CoV-2 vaccinations, there is a significant concern that CLD patients may be more susceptible to breakthrough infections and more severe outcomes than the overall population.^5–9^ A recent study utilizing the N3C Data Enclave showed that the estimated incidence rate for breakthrough infections in all fully vaccinated patients was approximately 5.0 per 1,000 person-months.^19^ In our study, we found that breakthrough infections occurred at a slightly higher rate for fully vaccinated CLD patients without cirrhosis (5.6 per 1,000 person-months) and at a similar rate for fully vaccinated CLD patients with cirrhosis (5.1 per 1,000 person-months). These suggest that CLD patients may not necessarily face a higher risk of breakthrough infections than the overall population. In multivariable incidence rate ratio calculations (Table 3), the presence of cirrhosis was not significantly associated with breakthrough infection. While this finding was initially surprising, it is congruent with antibody response studies of patients with and without cirrhosis to SARS-CoV-2 vaccinations, which showed that there were similar rates of neutralizing antibodies generated in CLD patients regardless of presence of cirrhosis.^8,9^

Consistent with other studies in CLD patient populations, we found that receiving the mRNA-1273 vaccine as the initial dose was associated with a 18% reduced risk for breakthrough infections compared with receiving the BNT162b2 vaccine.^16,26^ Moreover, we found a strong dose-dependent relationship between the number of vaccine doses received and decreased risks of breakthrough infection among CLD patients, consistent with revised Centers for Disease Control guidance around additional and “booster” shots given emergence of more transmissible and severe SARS-CoV-2 variants.^27^ While patients may not always have a choice as to the type of vaccination they could receive, these data could help inform recommendations for those who do have a choice between different types of vaccinations.

Most importantly, among CLD patients with cirrhosis and SARS-CoV-2 infection, vaccinated patients had lower rates of hospitalization, ventilation, and death within 30 days than unvaccinated patients. While CLD patients with cirrhosis and breakthrough infection could still have serious complications, full vaccination was associated with a 66% reduction in the risk of all-cause mortality within 30 days (aHR 0.34, 95% CI 0.17-0.68). This figure is notably less than the 78% reduction in all-cause mortality estimated from cohorts of veterans with CLD.^16,17^ Of particular note, partial vaccination was not statistically associated with a reduction in the risk of all-cause mortality in our analyses. As the point estimate suggested partial protection as expected from previous studies, we suspect that this may have been due to low power. Moreover, consistent with analyses of the N3C Data Enclave from earlier in the pandemic, the 30-day mortality rate for unvaccinated CLD patients with cirrhosis has remained at ∼9%, despite new treatment regimens and therapeutics. These findings are likely due to the impact of emerging SARS-CoV-2 variants, such as B.1.1.7 (Alpha) and B.1.617.2 (Delta),^28–30^ associated with more transmissible or severe disease.

While this study is one of the first on the impact of SARS-CoV-2 vaccinations in a large, gender-balanced, and diverse population of patients with CLD, there are several limitations that have been previously described in our works utilizing the N3C Data Enclave. Limitations due to the use of N3C Data Enclave include overrepresentation of tertiary academic medical centers, selection bias due to derivation of SARS-CoV-2 negative and positive populations based on testing, systematic missingness of certain variables due to data heterogeneity, and likely misclassification between patients with alcohol-associated liver disease and non-alcoholic fatty liver disease.^10^ These inherent limitations likely selected for a more clinically ill patient population than the general CLD and cirrhosis patient populations.

There are also several limitations specific to this particular study. First, the underlying vaccination data is based on recorded medication exposures at each of the data partner sites. This data may not include vaccinations administered at mass vaccination sites, local pharmacies, “pop-up” vaccination sites, and other venues. Therefore, there may be a significant proportion of false-negatives for detection of vaccine doses (patients who actually received a vaccination not recorded as such in the N3C Data Enclave). Second, while the N3C Data Enclave had additional or “booster” dose information in the recorded medication exposures at data partner sites, we were not able to estimate the mortality risk reduction associated an additional or “booster” dose due to the low number of death events occurring in this population (less than 20).^1^ We therefore consolidated those who received three or more vaccine doses into the full vaccination category in our analyses for our second study question. The impact of additional or “booster” shots on CLD patients will be an active area of investigation for us as N3C accumulates and harmonizes additional vaccination data.

Finally, our use of the de-identified version of the N3C Data Enclave hindered our ability to differentiate the impact of various SARS-CoV-2 variants. To protect patient privacy, date shifting was uniformly applied within each data site. This means that our analyses could not investigate temporal trends with each surge with accuracy. Based on the date on which we accessed the N3C Data Enclave (January 15, 2022) and our exclusion of patients whose date of SARS-CoV-2 testing was within 90 days of the calculated maximum data date for each site, our data likely reflects the B1.1.7 (Alpha) and B.1.617.2 (Delta) surges in the United States,^29,30^ but not the B.1.1.529 (Omicron) surge at the end of 2021 and beginning of 2022.^28,31^ The is a major limitation of our study and one that we hope to rectify in future analyses as the pandemic continues to evolve.

Despite this, our study is one of the largest studies of breakthrough infections and clinical outcomes in vaccinated CLD patients with and without cirrhosis. While generally consistent with results from other studies, our findings showed that CLD patients may not experience breakthrough infections at a higher rate than the overall population and that receiving an initial dose of mRNA-1273 vaccine may confer a marginally higher protection against breakthrough infections. In addition, we found partial vaccination was not significantly associated with mortality reduction, while full vaccination was associated with a 66% risk reduction. As vaccine hesitancy has been demonstrated in various clinical and demographic subsets of this patient population,^32^ we hope that this information will help providers to encourage vaccine uptake in CLD patients.

## Abbreviations

AALD: alcohol-associated liver disease
aHR: adjusted hazard ratio
BMI: body mass index
CDC: Centers for Disease Control and Prevention
CI: confidence interval
CLD: chronic liver disease
EHR: electronic health record
HR: hazard ratio
INR: international normalized ratio
IQR: interquartile range
IRB: institutional review board
IRR: incidence rate ratio
MELD-Na: Model for End-Stage Liver Disease-Sodium
mRNA: messenger ribonucleic acid
N3C: National COVID Cohort Collaborative
NAFLD: non-alcoholic fatty liver disease
NCATS: National Center for Advancing Translational Sciences
NIH: National Institutes of Health
NREVSS: National Respiratory and Enteric Virus Surveillance System
OMOP: Observational Medical Outcomes Partnership
SARS-CoV-2: Severe Acute Respiratory Syndrome Coronavirus 2
SE: standard error

## N3C Consortium Collaborators/Authors

N3C Consortium collaborators and authors are in the process of being documented.

## Financial Support

The authors of this study were supported by AASLD Anna S. Lok Advanced/Transplant Hepatology Award (AASLD Foundation, Ge), P30DK026743 (UCSF Liver Center Grant, Ge and Lai), F31HL156498 (National Heart, Lung, and Blood Institute, Digitale), UL1TR001872 (National Center for Advancing Translational Sciences, Pletcher), and R01AG059183 (National Institute on Aging, Lai).

The content is solely the responsibility of the authors and does not necessarily represent the official views of the National Institutes of Health or any other funding agencies. The funding agencies played no role in the analysis of the data or the preparation of this manuscript.

## Acknowledgements

The analyses described in this publication were conducted with data or tools accessed through the NCATS N3C Data Enclave covid.cd2h.org/enclave and supported by NCATS U24 TR002306. This research was possible because of the patients whose information is included within the data from participating organizations (covid.cd2h.org/dtas) and the organizations and scientists (covid.cd2h.org/duas) who have contributed to the on-going development of this community resource.^1^

The N3C data transfer to NCATS is performed under a Johns Hopkins University Reliance Protocol # IRB00249128 or individual site agreements with NIH. The N3C Data Enclave is managed under the authority of the NIH; information can be found at https://ncats.nih.gov/n3c/resources.

We gratefully acknowledge contributions from the following N3C core teams (Asterisks indicate leads):

- Principal Investigators: Melissa A. Haendel*, Christopher G. Chute*, Kenneth R. Gersing, Anita Walden
- Workstream, subgroup and administrative leaders: Melissa A. Haendel*, Tellen D. Bennett, Christopher G. Chute, David A. Eichmann, Justin Guinney, Warren A. Kibbe, Hongfang Liu, Philip R.O. Payne, Emily R. Pfaff, Peter N. Robinson, Joel H. Saltz, Heidi Spratt, Justin Starren, Christine Suver, Adam B. Wilcox, Andrew E. Williams, Chunlei Wu
- Key liaisons at data partner sites
- Regulatory staff at data partner sites
- Individuals at the sites who are responsible for creating the datasets and submitting data to N3C, Data Ingest and Harmonization Team: Christopher G. Chute*, Emily R. Pfaff*, Davera Gabriel, Stephanie S. Hong, Kristin Kostka, Harold P. Lehmann, Richard A. Moffitt, Michele Morris, Matvey B. Palchuk, Xiaohan Tanner Zhang, Richard L. Zhu
- Phenotype Team (Individuals who create the scripts that the sites use to submit their data, based on the COVID and Long COVID definitions): Emily R. Pfaff*, Benjamin Amor, Mark M. Bissell, Marshall Clark, Andrew T. Girvin, Stephanie S. Hong, Kristin Kostka, Adam M. Lee, Robert T. Miller, Michele Morris, Matvey B. Palchuk, Kellie M. Walters
- Project Management and Operations Team: Anita Walden*, Yooree Chae, Connor Cook, Alexandra Dest, Racquel R. Dietz, Thomas Dillon, Patricia A. Francis, Rafael Fuentes, Alexis Graves, Julie A. McMurry, Andrew J. Neumann, Shawn T. O’Neil, Usman Sheikh, Andréa M. Volz, Elizabeth Zampino
- Partners from NIH and other federal agencies: Christopher P. Austin*, Kenneth R. Gersing*, Samuel Bozzette, Mariam Deacy, Nicole Garbarini, Michael G. Kurilla, Sam G. Michael, Joni L. Rutter, Meredith Temple-O’Connor
- Analytics Team (Individuals who build the Enclave infrastructure, help create codesets, variables, and help Domain Teams and project teams with their datasets): Benjamin Amor*, Mark M. Bissell, Katie Rebecca Bradwell, Andrew T. Girvin, Amin Manna, Nabeel Qureshi
- Publication Committee Management Team: Mary Morrison Saltz*, Christine Suver*, Christopher G. Chute, Melissa A. Haendel, Julie A. McMurry, Andréa M. Volz, Anita Walden

The authors thank the Publication Committee for their review of this publication to ensure compliance with ICMJE guidelines, the N3C User Code of Conduct, and appropriate author attribution.

- Publication Committee Review Team: Carolyn Bramante, Jeremy R. Harper, Wendy Hernandez, Farrukh M. Koraishy, Saidulu Mattapally, Amit Saha, Satyanarayana Vedula

## Data Partners with Released Data

Stony Brook University — U24TR002306, University of Oklahoma Health Sciences Center — U54GM104938: Oklahoma Clinical and Translational Science Institute (OCTSI), West Virginia University — U54GM104942: West Virginia Clinical and Translational Science Institute (WVCTSI), University of Mississippi Medical Center — U54GM115428: Mississippi Center for Clinical and Translational Research (CCTR), University of Nebraska Medical Center — U54GM115458: Great Plains IDeA-Clinical & Translational Research, Maine Medical Center — U54GM115516: Northern New England Clinical & Translational Research (NNE-CTR) Network, Wake Forest University Health Sciences — UL1TR001420: Wake Forest Clinical and Translational Science Institute, Northwestern University at Chicago — UL1TR001422: Northwestern University Clinical and Translational Science Institute (NUCATS), University of Cincinnati — UL1TR001425: Center for Clinical and Translational Science and Training, The University of Texas Medical Branch at Galveston — UL1TR001439: The Institute for Translational Sciences, Medical University of South Carolina — UL1TR001450: South Carolina Clinical & Translational Research Institute (SCTR), University of Massachusetts Medical School Worcester — UL1TR001453: The UMass Center for Clinical and Translational Science (UMCCTS), University of Southern California — UL1TR001855: The Southern California Clinical and Translational Science Institute (SC CTSI), Columbia University Irving Medical Center — UL1TR001873: Irving Institute for Clinical and Translational Research, George Washington Children’s Research Institute — UL1TR001876: Clinical and Translational Science Institute at Children’s National (CTSA-CN), University of Kentucky — UL1TR001998: UK Center for Clinical and Translational Science, University of Rochester — UL1TR002001: UR Clinical & Translational Science Institute, University of Illinois at Chicago — UL1TR002003: UIC Center for Clinical and Translational Science, Penn State Health Milton S. Hershey Medical Center — UL1TR002014: Penn State Clinical and Translational Science Institute, The University of Michigan at Ann Arbor — UL1TR002240: Michigan Institute for Clinical and Health Research, Vanderbilt University Medical Center — UL1TR002243: Vanderbilt Institute for Clinical and Translational Research, University of Washington — UL1TR002319: Institute of Translational Health Sciences, Washington University in St. Louis — UL1TR002345: Institute of Clinical and Translational Sciences, Oregon Health & Science University — UL1TR002369: Oregon Clinical and Translational Research Institute, University of Wisconsin-Madison — UL1TR002373: UW Institute for Clinical and Translational Research, Rush University Medical Center — UL1TR002389: The Institute for Translational Medicine (ITM), The University of Chicago — UL1TR002389: The Institute for Translational Medicine (ITM), University of North Carolina at Chapel Hill — UL1TR002489: North Carolina Translational and Clinical Science Institute, University of Minnesota — UL1TR002494: Clinical and Translational Science Institute, Children’s Hospital Colorado — UL1TR002535: Colorado Clinical and Translational Sciences Institute, The University of Iowa — UL1TR002537: Institute for Clinical and Translational Science, The University of Utah — UL1TR002538: Uhealth Center for Clinical and Translational Science, Tufts Medical Center — UL1TR002544: Tufts Clinical and Translational Science Institute, Duke University — UL1TR002553: Duke Clinical and Translational Science Institute, Virginia Commonwealth University — UL1TR002649: C. Kenneth and Dianne Wright Center for Clinical and Translational Research, The Ohio State University — UL1TR002733: Center for Clinical and Translational Science, The University of Miami Leonard M. Miller School of Medicine — UL1TR002736: University of Miami Clinical and Translational Science Institute, University of Virginia — UL1TR003015: iTHRIV Integrated Translational health Research Institute of Virginia, Carilion Clinic — UL1TR003015: iTHRIV Integrated Translational health Research Institute of Virginia, University of Alabama at Birmingham — UL1TR003096: Center for Clinical and Translational Science, Johns Hopkins University — UL1TR003098: Johns Hopkins Institute for Clinical and Translational Research, University of Arkansas for Medical Sciences — UL1TR003107: UAMS Translational Research Institute, Nemours — U54GM104941: Delaware CTR ACCEL Program, University Medical Center New Orleans — U54GM104940: Louisiana Clinical and Translational Science (LA CaTS) Center, University of Colorado Denver, Anschutz Medical Campus — UL1TR002535: Colorado Clinical and Translational Sciences Institute, Mayo Clinic Rochester — UL1TR002377: Mayo Clinic Center for Clinical and Translational Science (CCaTS), Tulane University — UL1TR003096: Center for Clinical and Translational Science, Loyola University Medical Center — UL1TR002389: The Institute for Translational Medicine (ITM), Advocate Health Care Network — UL1TR002389: The Institute for Translational Medicine (ITM), OCHIN — INV-018455: Bill and Melinda Gates Foundation grant to Sage Bionetworks

## Additional Data Partners Who Have Signed a DTA and Whose Data Release is Pending

The Rockefeller University — UL1TR001866: Center for Clinical and Translational Science, The Scripps Research Institute — UL1TR002550: Scripps Research Translational Institute, University of Texas Health Science Center at San Antonio — UL1TR002645: Institute for Integration of Medicine and Science, The University of Texas Health Science Center at Houston — UL1TR003167: Center for Clinical and Translational Sciences (CCTS), NorthShore University HealthSystem — UL1TR002389: The Institute for Translational Medicine (ITM), Yale New Haven Hospital — UL1TR001863: Yale Center for Clinical Investigation, Emory University — UL1TR002378: Georgia Clinical and Translational Science Alliance, Weill Medical College of Cornell University — UL1TR002384: Weill Cornell Medicine Clinical and Translational Science Center, Montefiore Medical Center — UL1TR002556: Institute for Clinical and Translational Research at Einstein and Montefiore, Medical College of Wisconsin — UL1TR001436: Clinical and Translational Science Institute of Southeast Wisconsin, University of New Mexico Health Sciences Center — UL1TR001449: University of New Mexico Clinical and Translational Science Center, George Washington University — UL1TR001876: Clinical and Translational Science Institute at Children’s National (CTSA-CN), Stanford University — UL1TR003142: Spectrum: The Stanford Center for Clinical and Translational Research and Education, Regenstrief Institute — UL1TR002529: Indiana Clinical and Translational Science Institute, Cincinnati Children’s Hospital Medical Center — UL1TR001425: Center for Clinical and Translational Science and Training, Boston University Medical Campus — UL1TR001430: Boston University Clinical and Translational Science Institute, The State University of New York at Buffalo — UL1TR001412: Clinical and Translational Science Institute, Aurora Health Care — UL1TR002373: Wisconsin Network For Health Research, Brown University — U54GM115677: Advance Clinical Translational Research (Advance-CTR), Rutgers, The State University of New Jersey — UL1TR003017: New Jersey Alliance for Clinical and Translational Science, Loyola University Chicago — UL1TR002389: The Institute for Translational Medicine (ITM), #N/A — UL1TR001445: Langone Health’s Clinical and Translational Science Institute, Children’s Hospital of Philadelphia — UL1TR001878: Institute for Translational Medicine and Therapeutics, University of Kansas Medical Center — UL1TR002366: Frontiers: University of Kansas Clinical and Translational Science Institute, Massachusetts General Brigham — UL1TR002541: Harvard Catalyst, Icahn School of Medicine at Mount Sinai — UL1TR001433: ConduITS Institute for Translational Sciences, Ochsner Medical Center — U54GM104940: Louisiana Clinical and Translational Science (LA CaTS) Center, HonorHealth — None (Voluntary), University of California, Irvine — UL1TR001414: The UC Irvine Institute for Clinical and Translational Science (ICTS), University of California, San Diego — UL1TR001442: Altman Clinical and Translational Research Institute, University of California, Davis — UL1TR001860: UCDavis Health Clinical and Translational Science Center, University of California, San Francisco — UL1TR001872: UCSF Clinical and Translational Science Institute, University of California, Los Angeles — UL1TR001881: UCLA Clinical Translational Science Institute, University of Vermont — U54GM115516: Northern New England Clinical & Translational Research (NNE-CTR) Network, Arkansas Children’s Hospital — UL1TR003107: UAMS Translational Research Institute

## Author Contributions

Authorship was determined using ICMJE recommendations.

*Ge*: Study concept and design; data extraction; analysis and interpretation of data; drafting of manuscript; critical revision of the manuscript for important intellectual content; statistical analysis

*Digitale*: Analysis and interpretation of data; drafting of manuscript; critical revision of the manuscript for important intellectual content

*Pletcher*: Acquisition of data; interpretation of data; critical revision of the manuscript for important intellectual content

*Lai*: Study concept and design; analysis and interpretation of data; drafting of manuscript; critical revision of the manuscript for important intellectual content; obtained funding; study supervision

## N3C Consortium Collaborators/Authors Contributions

To be populated based on input from N3C Publications Committee

## Conflicts of Interest

The authors have no conflicts of interest to declare.

## Writing Assistance

None.

## Data Availability Statement

The N3C Data Enclave (covid.cd2h.org/enclave) houses fully reproducible, transparent, and broadly available limited and de-identified datasets (HIPAA definitions: https://www.hhs.gov/hipaa/for-professionals/privacy/specialtopics/de-identification/index.html). Data is accessible by investigators at institutions that have signed a Data Use Agreement with NIH who have taken human subjects and security training and attest to the N3C User Code of Conduct. Investigators wishing to access the limited dataset must also supply an institutional IRB protocol. All requests for data access are reviewed by the NIH Data Access Committee. A full description of the N3C Enclave governance has been published; information about how to apply for access is available on the NCATS website: https://ncats.nih.gov/n3c/about/applying-for-access. Reviewers and health authorities will be given access permission and guidance to aid reproducibility and outcomes assessment. A Frequently Asked Questions about the data and access has been created at: https://ncats.nih.gov/n3c/about/program-faq The data model is OMOP 5.3.1, specifications are posted at: https://ncats.nih.gov/files/OMOP_CDM_COVID.pdf

## Notes

### Competing Interest Statement

The authors have declared no competing interest.

### Summary of Updates

Minor calculation changes.

