## Supplemental Table 1 for "Breakthrough SARS-CoV-2 Infection Outcomes in Vaccinated Patients with Chronic Liver Disease and Cirrhosis: A National COVID Cohort Collaborative Study"

**Supplemental Table 1 – Standard OMOP concept identifiers for SARS-CoV-2 vaccinations**

| **OMOP Concept Identifier(s)** | **Concept Name(s)** | **Corresponding Vaccination** |
| --- | --- | --- |
| 739906 | SARS-COV-2 (COVID-19) vaccine, vector - Ad26 100000000000 UNT/ML Injectable Suspension | JNJ-784336725 |
| 739903 | SARS-COV-2 (COVID-19) vaccine, vector - Ad26 100000000000 UNT/ML | JNJ-784336725 |
| 702866 | SARS-COV-2 (COVID-19) vaccine, vector non-replicating, recombinant spike protein-Ad26, preservative free, 0.5 mL | JNJ-784336725 |
| 37003436 | SARS-CoV-2 (COVID-19) vaccine, mRNA-BNT162b2 0.1 MG/ML Injectable Suspension | BNT162b2 |
| 37003517 | SARS-CoV-2 (COVID-19) vaccine, mRNA-1273 0.2 MG/ML | mRNA-1273 |
| 37003433 | SARS-CoV-2 (COVID-19) vaccine, mRNA-BNT162b2 0.1 MG/ML | BNT162b2 |
| 37003518 | SARS-CoV-2 (COVID-19) vaccine, mRNA-1273 0.2 MG/ML Injectable Suspension | mRNA-1273 |
